# Systematic review protocol exploring the impact of the COVID-19 pandemic on the wellbeing of general practitioners

**DOI:** 10.1101/2021.08.05.21261627

**Authors:** L. Jefferson, S. Golder, V. Dale, H. Essex, E. McHugh, K. Bloor

## Abstract

**Background:** Over recent years chronic stress and burnout have been reported by doctors working in general practice in the UK NHS and internationally. The COVID-19 pandemic has changed general practitioners’ working lives – adding potential pressures from avoiding infection and addressing pent-up demand for care, but also changing processes such as rapidly taking up remote consultations. To date, there has been a focus on exploring the impact of the pandemic on the wellbeing of hospital clinicians. No registered systematic reviews currently focus on exploring the impact of the pandemic on the mental health and wellbeing of general practitioners.

**Aims and objectives:** To synthesise the current international evidence base exploring the impact of COVID-19 on the mental health and wellbeing of general practitioners, and which factors are associated with their reported mental health and wellbeing during the pandemic.

**Methods:** In this paper we report a systematic review protocol, following PRISMA guidance. In our search strategy we will identify primary research studies or systematic reviews exploring the mental health and wellbeing of general practitioners during the COVID-19 pandemic in four databases (MEDLINE, Embase, PsychInfo and Medrxiv) and Google Scholar. We will hand-search reference lists and grey literature.

Two reviewers will undertake all stages including study selection, data extraction and quality assessment, with arbitration by a third reviewer where necessary. We will use standardised quality assessment tools to ensure transparency and reduce bias in quality assessment. Depending on the quality of included studies, we may undertake a sensitivity analysis by excluding studies from narrative synthesis that are rated as low quality using the checklists.

We will describe the findings across studies using narrative thematic data synthesis, and if sufficiently homogenous data are identified, we will pool quantitative findings through meta-analysis.

## Introduction

Prior to the COVID-19 pandemic, chronic stress and burnout threatened the mental health of doctors working in general practice, the quality of patient care^1^ and the sustainability of the health care system.^2^ General practitioners (GPs) play a crucial role as ‘gatekeepers’ to the health service, and yet greater quantity and complexity of work have led to high levels of reported job stress, lower satisfaction, burnout and intentions to leave medicine.^3,4,5,6^

Internationally, medical workforce wellbeing has been described as a “public health crisis.”^7,8^ In the UK, prior to the pandemic, 80% of doctors reported characteristics associated with very high risk of burnout,^9^ with those in general practice and emergency medicine at highest risk.^10,11^

There are clear added risks to the wellbeing of the medical workforce during COVID-19, and during the recovery period, as we emerge from the pandemic with a large backlog of unmet patient needs for non-COVID-19 care. Research from earlier epidemics indicates the potential for considerable increased stress and burnout in health professionals.^12^ Sources of stress for GPs include rapid change, risks of infection, remote working and reductions in face-to-face patient care. There may also, however, be some changes that could improve GPs’ wellbeing, including public gratitude, financial investment, e-consultations and improved cooperation; factors previously shown to positively affect workforce morale and wellbeing.

Internationally, research evidence tends to focus on the impact of the COVID-19 pandemic on front-line hospital workers, perhaps due to their roles in treating the illness and potential greater exposure to infection. A systematic review of international evidence is needed to explore the potential impact of COVID-19 on the mental health and wellbeing of doctors working in general practice, so that future health policy and workplace interventions can be tailored to suit these specific experiences.

## Aims

In this systematic review we aim to synthesise current international evidence on the impact of COVID-19 on the mental health and wellbeing of general practitioners. In addition, and if data allows, we will explore factors which are associated with the mental health and wellbeing of general practitioners during the COVID-19 pandemic.

## Methods

An initial scoping exercise, searching PROSPERO-registered studies and contacting study authors to avoid duplication in this rapidly expanding research field, revealed no existing reviews with a focus on general practitioner mental health and wellbeing during the COVID-19 pandemic.

To ensure the transparency of reporting for this systematic review we will use the Preferred Reporting Items for Systematic Reviews and Meta-Analyses (PRISMA) guidance.^13^ Our eligibility criteria will capture studies with GPs as the population of interest, which measure mental health and wellbeing outcomes during the COVID-19 pandemic.

### Criteria for including studies

#### Population

General practitioners. We will exclude studies of doctors not working in primary care. We will exclude studies of a variety of health professionals, including GPs, if they do not provide subgroup analyses or data specifically for the GP group.

#### Intervention / type of exposure

the impact of the COVID-19 pandemic.

#### Comparator

This is not an interventional systematic review; therefore, no comparator will be used. Studies may explore trends over time.

#### Outcome measures

Measures of psychological wellbeing, stress and burnout, and as secondary outcomes, the impact on factors including absenteeism and workforce retention.

#### Study Design

Studies will not be limited by design, but we will include only empirical primary research and systematic reviews. Editorials and case reports will be excluded.

### Search methods for identification of studies

We will develop a search strategy with input from a multi-disciplinary research team, including subject matter experts, an experienced information specialist and through consultation with GP and patient stakeholders. We will search the following databases; MEDLINE, Embase, PsychInfo and, owing to the large body of COVID-19 literature being published in this space, Medrxiv and Google Scholar will also be searched. We will search from 2020 onwards and limit to the English language, using search terms for general practitioner and relating to general practice, including consideration of international terminology, for example the inclusion of terms relating to ‘family practice.’ Search terms will focus on COVID-19, as well as date restrictions to only include studies published during 2020 and 2021. We will hand search reference lists and grey literature. We will update searches prior to publication to ensure inclusion of all literature in this rapidly expanding field.

#### Study selection

We will download all records to Endnote, removing any duplicated studies. Two reviewers will independently screen studies for inclusion.

In the first stage of screening, we will classify all titles and abstracts as eligible, excluded or uncertain about eligibility. Each reviewer will be blind to the other’s assessment. The full texts for all eligible or potentially eligible papers will then be retrieved and reviewed. A third reviewer will resolve any disagreements regarding inclusion of studies.

We will record excluded articles, with the reasons for exclusion, to generate a flow diagram as recommended by the PRISMA statement.^14^

#### Data extraction

A data extraction form will be designed and piloted including information regarding study design, sample size, sample characteristics, primary and secondary outcomes. One reviewer will perform data extraction, with a 20% sample second checked by a second reviewer.

#### Assessment of methodological quality

We will assess the quality of identified studies using standardised checklists, chosen according to the study design. We will use the AMSTAR checklist^15^ for reviews, the CASP quality checklist^16^ for observational and qualitative studies, and the Joanna Briggs Institute checklist for Analytical Cross-sectional Studies.^17^ Depending on the quality of included studies, we may undertake a sensitivity analysis by excluding studies from narrative synthesis that are rated as low quality using the checklists. Two reviewers will independently conduct quality appraisal, with disagreements resolved by a third reviewer.

#### Synthesis of findings

Our data synthesis will be narrative, using a thematic approach to describe the findings across studies, with subgroup analyses according to study type. Where appropriate (i.e. sufficiently homogenous data and study type), studies with quantitative findings will be pooled using meta-analysis. We will analyse subgroups of study types. Where sufficient data is available, we will analyse different subgroups of key GP characteristics (such as career stage, gender, ethnicity).

#### Protocol amendments

Where changes are required to this protocol, the details will be outlined in the published final review and updated in PROSPERO. However, no further amendments to this protocol are foreseen.

## Discussion

Internationally, general practitioners report increasing stress and burnout, with clear potential additional pressures likely during the COVID-19 pandemic and recovery period. There are currently no systematic reviews of general practitioners’ wellbeing during the COVID-19 pandemic. Given the complexity and quantity of workload stressors placed on general practitioners prior to and during the pandemic, this review is timely in order to highlight how best to support the future working lives of GPs. In this study we aim to synthesise this international evidence base to explore the impact of COVID-19 on the mental health and wellbeing of general practitioners, which will inform future workforce policy as we move out of the pandemic. We anticipate this work will also inform future research, including the potential to design and implement workplace interventions to support the wellbeing of general practice teams. We will disseminate findings from this systematic review through peer-reviewed journals and presentations to national audiences including policymakers and the Royal College of General Practitioners.

## Data Availability

This is a protocol for a systematic review, as such no data is currently available.

## Author contributions

This study was designed and conceived by LJ and KB, with methodological expertise input from SG. LJ wrote the first draft of this manuscript, to which all authors commented. All authors have read and agreed the final version.

